# Association of Vitamin D Deficiency and Treatment with COVID-19 Incidence

**DOI:** 10.1101/2020.05.08.20095893

**Authors:** David O. Meltzer, Thomas J. Best, Hui Zhang, Tamara Vokes, Vineet Arora, Julian Solway

## Abstract

**Importance:** Vitamin D treatment has been found to decrease incidence of viral respiratory tract infection, especially in vitamin D deficiency. It is unknown whether COVID-19 incidence is associated with vitamin D deficiency and treatment.

**Objective:** To examine whether vitamin D deficiency and treatment are associated with testing positive for COVID-19.

**Design:** Retrospective cohort study

**Setting:** University of Chicago Medicine

**Participants:** Patients tested for COVID-19 from 3/3/2020-4/10/2020. Vitamin D deficiency was defined by the most recent 25-hydroxycholecalciferol <20ng/ml or 1,25-dihydroxycholecalciferol <18pg/ml within 1 year before COVID-19 testing. Treatment was defined by the most recent vitamin D type and dose, and treatment changes between the time of the most recent vitamin D level and time of COVID-19 testing. Vitamin D deficiency and treatment changes were combined to categorize vitamin D status at the time of COVID-19 testing as likely deficient (last-level-deficient/treatment-not-increased), likely sufficient(last-level-not-deficient/treatment-not-decreased), or uncertain deficiency(last-level-deficient/treatment-increased or last-level-not-deficient/treatment-decreased).

**Main Outcomes and Measures:** The main outcome was testing positive for COVID-19. Multivariable analysis tested whether the most recent vitamin D level and treatment changes after that level were associated with testing positive for COVID-19 controlling for demographic and comorbidity indicators. Bivariate analyses of associations of treatment with vitamin D deficiency and COVID-19 were performed.

**Results:** Among 4,314 patients tested for COVID-19, 499 had a vitamin D level in the year before testing. Vitamin D status at the time of COVID-19 testing was categorized as likely deficient for 127(25%) patients, likely sufficient for 291(58%) patients, and uncertain for 81(16%) patients. In multivariate analysis, testing positive for COVID-19 was associated with increasing age(RR(age<50)=1.05,p<0.021;RR(age≥50)=1.02,p<0.064)), non-white race(RR=2.54,p<0.01) and being likely vitamin D deficient (deficient/treatment-not-increased:RR=1.77,p<0.02) as compared to likely vitamin D sufficient(not-deficient/treatment-not-decreased), with predicted COVID-19 rates in the vitamin D deficient group of 21.6%(95%CI[14.0%-29.2%]) versus 12.2%(95%CI[8.9%-15.4%]) in the vitamin D sufficient group. Vitamin D deficiency declined with increasing vitamin D dose, especially of vitamin D3. Vitamin D dose was not significantly associated with testing positive for COVID-19.

**Conclusions and Relevance:** Vitamin D deficiency that is not sufficiently treated is associated with COVID-19 risk. Testing and treatment for vitamin D deficiency to address COVID-19 warrant aggressive pursuit and study.

---

COVID-19, caused by the SARS-CoV-2 coronavirus, often produces severe lower respiratory symptoms and has caused 200,000 deaths worldwide.^1^ One challenge in halting this pandemic is the absence of evidence demonstrating effective pharmacologic interventions to prevent COVID-19. Vitamin D has been identified as a potential strategy to prevent or treat COVID-19.^2^ Vitamin D treatment has been found to decrease other viral respiratory infections, especially in persons with vitamin D deficiency.^3^ Vitamin D deficiency is common, affecting nearly half the US population, with higher rates among persons with darker skin or reduced sun exposure, including persons living in higher latitudes in the winter, nursing home residents, and health care workers.^4^ COVID-19 is more prevalent among African-Americans,^5^ persons living in northern cities in the late winter,^6^ older adults,^7^ nursing home residents^8^ and health care workers, populations which all have increased risk of vitamin D deficiency.^10,11,12,13^ Moreover, COVID-19 is less prevalent in pregnant women and children^14^, and in persons living in Japan^15^, in whom rates of vitamin D deficiency are lower.^16,17,18^ Shelter in place orders to reduce the spread of COVID-19 may also decrease sun exposure, potentially worsening vitamin D levels and providing further rationale for vitamin D supplementation.^19^ Given the low risks and low cost of vitamin D treatment, it has been suggested that vitamin D treatment should be rapidly scaled based on existing evidence.^20,^ Nevertheless, evidence of whether vitamin D deficiency is associated with COVID-19 infection is lacking and could help establish vitamin D as an evidence-based approach to decrease the burden and potentially the spread of COVID-19.

Using data from the electronic health record at the University of Chicago Medicine (UCM) in Chicago, IL, we examined whether persons tested for COVID-19 were more likely to test positive for COVID-19 if they were vitamin D deficient than if they were not vitamin D deficient. Because patients may have had changes in their vitamin D treatment after the time of their most recent vitamin D level prior to COVID-19 testing, we combined data on patients’ last vitamin D level before COVID-19 testing and changes in their treatment after that vitamin D level to construct a measure of whether each patient was expected to have a deficient, sufficient, or uncertain vitamin D level at the time they were tested for COVID-19. We also tested whether the type and dosage of vitamin D supplementation was associated with vitamin D levels and COVID-19.

## Methods

### Study Design and Oversight

The underlying motivation for our analysis is that if vitamin D deficiency increases the risk of COVID-19, then persons with vitamin D deficiency tested for COVID-19 should be more likely to test positive than persons being tested who are not vitamin D deficient. Based on this hypothesis, we obtained IRB approval for a preliminary bivariate analysis examining whether patients tested for COVID-19 at UCM were more likely to test positive for COVID-19 if they had a documented deficient level of vitamin D in the past 2 years compared to patients with non-deficient vitamin D levels. This preliminary analysis demonstrated an increased rate of testing positive for COVID-19 among persons with vitamin D deficiency and suggested the importance of examining data on vitamin D treatments after the time of these test results. Since a large fraction of the persons tested at UCM were UCM health care workers, exploratory analyses were begun to inform UCM internal operations in parallel with IRB submission of this study, which was approved by the University of Chicago Biological Sciences Division IRB.

### Participants

We obtained data for all 4,314 persons tested for COVID-19 at UCM from March 3, 2020 to April 10, 2020. We obtained electronic health record data for required demographic, comorbidity, laboratory and medication data, including 25(OH)-vitamin D levels within 2 years before COVID-19 testing. Vitamin D levels and treatments within 14 days of COVID-19 testing were excluded from analyses to avoid possible confounding by potential early manifestations of COVID-19, such as presenting for health care with symptoms that could lead to testing for and treatment of vitamin D deficiency. This resulted in 22 patients being excluded.

### Measurements

All variables were defined based on information from the UCM electronic health record, Epic® (Verona, WI). COVID-19 test status was determined by any positive COVID-19 RNA test result. Because of limitations on testing, testing at UCM was focused on persons presenting with potential symptoms of COVID-19 admitted to the hospital or health care workers with COVID-19 symptoms and exposure. The first date of testing positive for COVID-19 was used to determine the testing date for calculating variables defined in relation to the testing date. Patients with a most recent 25(OH)-vitamin D level <20 ng/ml within 1 year of COVID-19 testing were deemed vitamin D deficient. Patients with a most recent vitamin D levels within 1 year ≥20 ng/ml were deemed not vitamin D deficient. Patients treated with calcitriol and with calcitriol levels were classified as vitamin D deficient if the calcitriol levels <18 pg/ml. Vitamin D3 dosing was defined based on most recent daily dose recorded over the past 2 years excluding the 14 days before testing positive: None, 1-1000IU or a multivitamin, 2000IU, ≥3000IU. Indicators for treatment with D2 and calcitriol were also included. To account for possible changes in patients’ vitamin D treatment after the time of their last vitamin D level, we combined the data on the last vitamin D level and changes in treatment after that vitamin D level to assign each patient to one of four categories reflecting their likelihood of being vitamin D deficient at the time of COVID-19 testing: 1) deficient/treatment-not-increased (likely deficient), 2) not-deficient/treatment-not-decreased (likely sufficient), and 3) deficient/treatment-increased and 4) not-deficient/treatment-decreased, indicating uncertain vitamin D sufficiency. Days from most recent vitamin D level to COVID-19 diagnosis was constructed for sensitivity analysis.

Age, gender, race, Hispanic ethnicity were also obtained and coded as reported in Table 1. BMI and the following ICD-10-CM-based Elixhauser comorbidity clusters^21^ potentially related to COVID-19 coded based on all encounters during the period: hypertension, diabetes, chronic pulmonary disease, pulmonary circulation disorders, depression, and immunosuppression. Comorbidity indicators for liver disease and chronic kidney disease were also included because their potential roles in affecting levels of activated vitamin D.

**Table 1:**
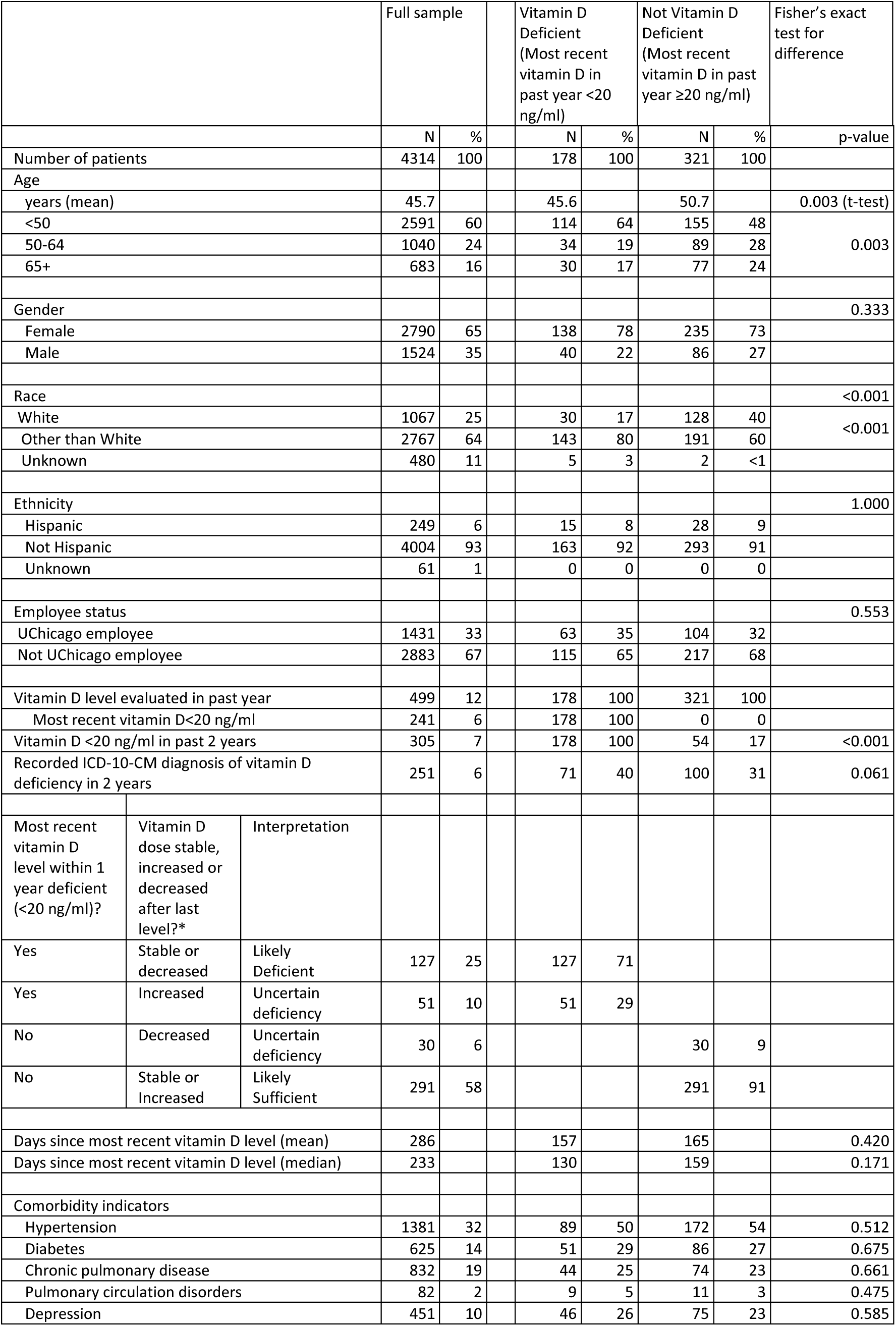
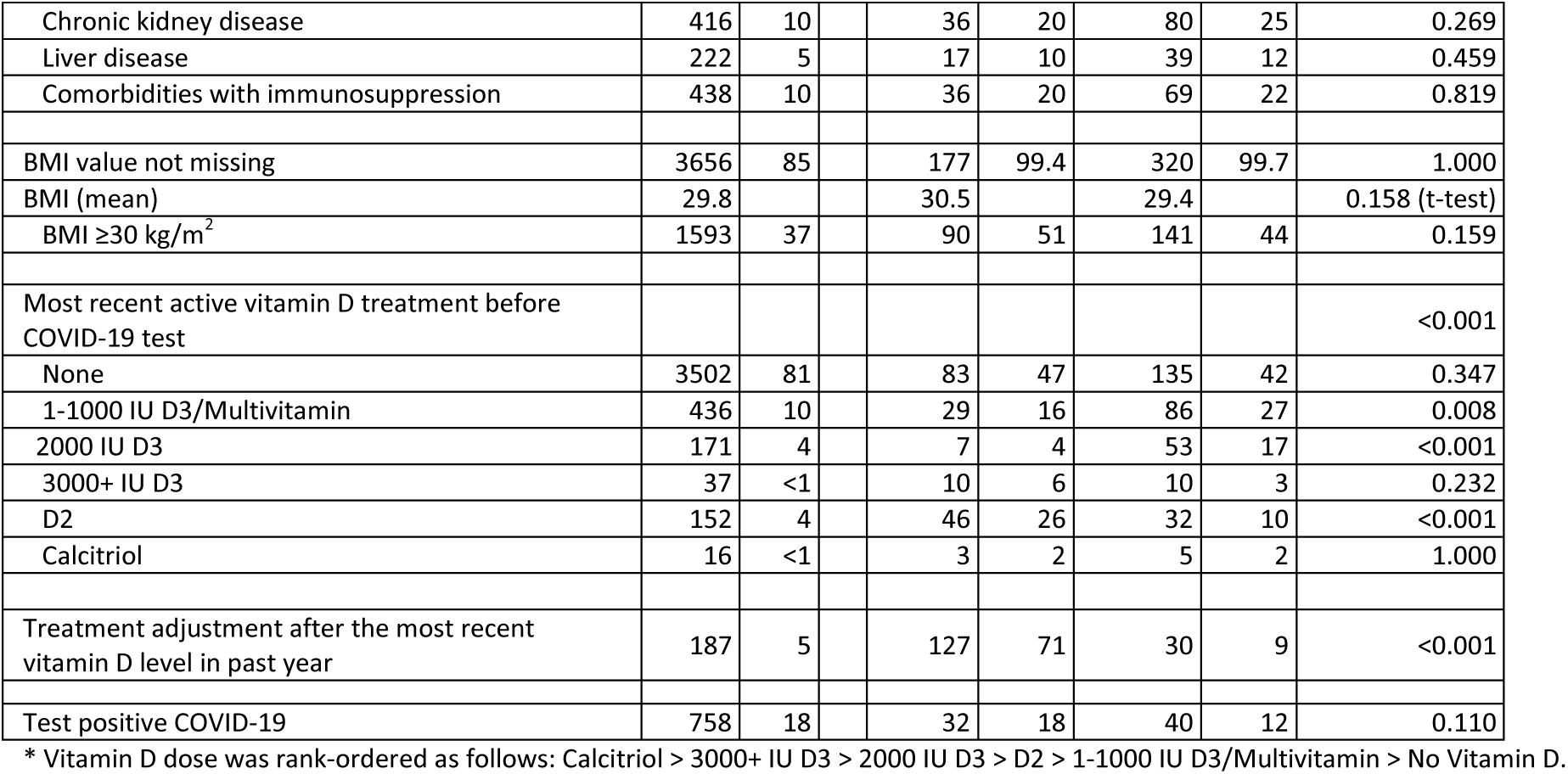
Characteristics of Patient Population

### Statistical Analysis

Basic descriptive statistics were reviewed for all variables. Fisher’s exact test was used to compare rates of testing positive for persons with last vitamin D levels that were deficient and persons with last levels that were not deficient. A multivariable generalized linear model (GLM) with binomial residuals and log link function^22^ was estimated with the covariates noted above. We also tested whether treatment dose/type was associated with vitamin D deficiency and COVID-19. In the bivariate COVID-19 analysis, patients were included if they had a diagnosis of vitamin D deficiency defined by having any deficient vitamin D level or recorded ICD-10-CM code for vitamin D deficiency within 2 years.

## Results

### Characteristics of the Patients

All 4,314 patients tested for COVID-19 during the study period had a test result. Of these, 499 had a vitamin D level in the year before testing, including 178(36%) with vitamin D deficiency. Four-hundred-twenty-eight(10%) patients tested for COVID-19 had a diagnosis of vitamin D deficiency within 2 years. Table 1 provides descriptive statistics for all patients with a COVID-19 test result and comparisons of demographic and comorbidity indicators, treatments and rates of testing positive for COVID-19 for the cohorts of patients stratified by whether their last vitamin D level was deficient.

Table 1 indicates no significant differences between patients whose last vitamin D level was or was not deficient except that vitamin D deficient patients are more likely to be younger, African American, have deficient vitamin D levels, and receive vitamin D2 and less likely to receive vitamin D3.

Thirty-two of 178(18%) vitamin D deficient patients tested positive for COVID-19 versus 40 of 321(11%) non-deficient patients(p=0.11).

Supplemental Table 1 repeats these descriptive analyses dividing the sample among the 4 groups combining deficiency status and treatment changes after the last vitamin D level to infer vitamin D status at the time of COVID-19 testing. The results are similar but show some additional differences in some baseline attributes in the small groups with uncertain deficiency status. Controlling for these variables or omitting the groups with uncertain vitamin D status at the time of COVID-19 testing from the analysis did not change our findings.

Figure 1 depicts the distribution of most recent vitamin D levels between 1 year before and 14 days before COVID-19 test orders for the sample used in multivariable analysis (Table 2).

**Figure 1.**
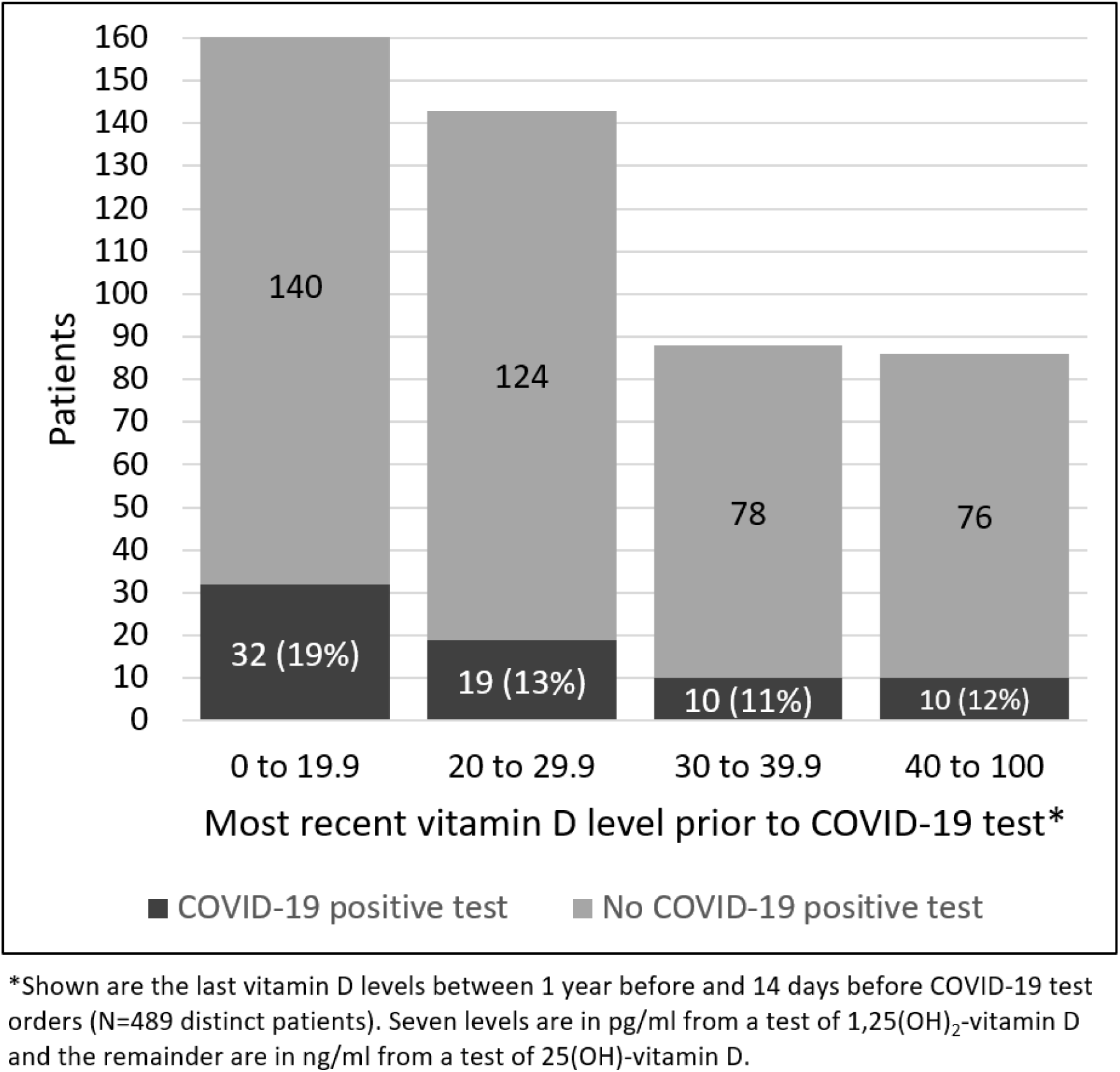
Most recent vitamin D levels prior to COVID-19 tests

**Table 2.** Multivariable Association of Vitamin D Deficiency and Treatment with Testing Positive for COVID-19

|  |  |  | N | % | Relative Risk | p-value |
| --- | --- | --- | --- | --- | --- | --- |
| Number of patients |  |  | 489 | 100 |  |  |
| Age (Linear Spline)<sup>§</sup> |  |  |  |  |  |  |
| <50 |  |  | 260 | 53 | 1.05 | 0.020 |
| 50+ |  |  | 229 | 47 | 1.02 | 0.063 |
| Gender (Male is reference) |  |  | 123 | 25 |  |  |
| Female |  |  | 366 | 75 | 0.87 | 0.578 |
| Race (White is reference) |  |  | 158 | 32 |  |  |
| Race other than white |  |  | 331 | 68 | 2.54 | 0.009 |
| Ethnicity (Not Hispanic is reference) |  |  | 448 | 92 |  |  |
| Hispanic |  |  | 41 | 8 | 0.29 | 0.209 |
| UChicago employee status (Not employee is reference) |  |  | 328 | 67 |  |  |
| UChicago Employee |  |  | 161 | 33 | 0.93 | 0.789 |
| Most recent vitamin D level within 1 year deficient (<20 ng/ml)? | Vitamin D dose stable, increased or decreased after last level?* | Interpretation |  |  |  |  |
| Yes | Stable or decreased* | Likely deficient | 124 | 27 | 1.77 | 0.015 |
| Yes | Increased | Uncertain deficiency | 48 | 10 | 1.10 | 0.823 |
| No | Decreased | Uncertain deficiency | 30 | 5 | 1.09 | 0.851 |
| No | Stable or Increased | Likely sufficient | 287 | 59 | reference |  |
| Comorbidity indicators |  |  |  |  |  |  |
| Hypertension |  |  | 261 | 53 | 1.08 | 0.789 |
| Diabetes |  |  | 137 | 28 | 0.78 | 0.314 |
| Chronic pulmonary disease |  |  | 117 | 24 | 0.91 | 0.727 |
| Pulmonary circulation disorders |  |  | 20 | 4 | 0.64 | 0.398 |
| Depression |  |  | 119 | 24 | 1.22 | 0.442 |
| Chronic kidney disease |  |  | 116 | 24 | 0.80 | 0.388 |
| Liver disease |  |  | 56 | 11 | 0.99 | 0.983 |
| Comorbidities with immunosuppression |  |  | 105 | 21 | 0.39 | 0.005 |
| BMI (mean) |  |  | 29.8 |  | 1.02 | 0.098 |
| Goodness-of-link test of squared predicted value<sup>30</sup> |  |  |  |  |  | 0.866 |
| Hosmer-Lemeshow goodness-of-fit decile test<sup>31</sup> |  |  |  |  |  | 0.889 |

§ A piecewise linear spline with a single knot at 50 improved model fit over models with unadjusted age or more complex parameterizations.
\* Vitamin D dose was rank-ordered as follows: Calcitriol > 3000+ IU D3 > 2000 IU D3 > D2 > 1-1000 IU D3/Multivitamin > No Vitamin D.

Among the 35% of patients whose most recent vitamin D-level was deficient, 18% tested positive for COVID-19, compared to 12% for patients whose last vitamin D level was not deficient(p=0.11).

### Follow-up and Outcomes

Table 2 shows the results of the multivariable GLM model for testing positive for COVID-19. Patients who were likely vitamin D deficient at the time of COVID-19 testing (i.e., their most recent vitamin D level before COVID-19 testing was deficient and they did not have their vitamin D dose increased) had an increased relative risk of testing positive for COVID-19(RR=1.77,p<0.02) compared to patients whose vitamin D level at the time of COVID-19 testing was likely sufficient (i.e., their last vitamin D level was not deficient and they at least maintained their vitamin D dose between the time of their last vitamin D level and when they were tested for COVID-19), for an estimated average rate in the vitamin D deficient group of 21.6%(95%CI[14.0%-29.2%]) versus 12.2%(95%CI[8.9%-15.4%]) in the vitamin D sufficient group. Testing positive for COVID-19 was also associated with increasing age, and non-white race, and was not associated with comorbidities except for a decreased incidence in persons with conditions associated with immunosuppression. Testing positive for COVID-19 was not significantly more likely for the patients classified as having vitamin D levels of uncertain sufficiency compared to patients with sufficient vitamin D levels. The risk of testing positive for COVID-19 in the likely deficient vitamin D group with a deficient last vitamin D level and no increase in treatment after that level was also not statistically significantly different than the risk of testing positive in the group with a deficient last vitamin D level but some increase in treatment. All these results were robust to including measures of time from last vitamin D level to COVID-19 testing. They also were preserved when the analysis was performed separately for non-white and white persons, though the small number of white persons forced dropping some covariates for model estimation in that subgroup. Because hypertension, obesity and diabetes may be ameliorated by treatment with Vitamin D, sensitivity analyses were also performed omitting these as covariates; this did not significantly change these findings.

Among the 124 patients who had a deficient last vitamin D level and did not have vitamin D treatment increased, 60% had no recorded vitamin D treatment after the deficient level, 30% continued their dosing before the deficient vitamin D level, and the remaining 10% primarily reflected switches from vitamin D3 to vitamin D2 or from vitamin D2 to ≤1000IU vitamin D3. Among patients with a deficient last vitamin D level who had an increase in treatment after that level, 35% initiated or increased vitamin D3 to 2000IU or more, 50% initiated D2, and 15% initiated low-dose ≤1000 IU vitamin D3.

Vitamin D treatment type and dosage were not significantly associated with testing positive for COVID-19 (Table 3) but the analysis was notable for a small number of individuals receiving higher doses of vitamin D3. Vitamin D type and dosage were significantly (p<0.01) associated with deficient vitamin D levels (Table 4). The latter effect was no longer significant (p=0.24) if patients in the D3 2000IU and ≥3000IU categories were omitted from the analysis.

**Table 3.** Bivariate analysis of most recent active vitamin D treatment 14 days before COVID-19 test order and COVID-19 test results, among patients with a diagnosis of vitamin D deficiency within 2 years or any level <20 ng/ml.

| Most recent active vitamin D treatment prior to COVID-19 test order | Total patients | COVID-19 positive |  |
| --- | --- | --- | --- |
|  |  | N | % |
| No Vitamin D | 180 | 23 | 13 |
| 1-1000 IU D3/Multivitamin | 88 | 18 | 20 |
| 2000 IU D3 | 41 | 6 | 15 |
| 3000+ IU D3 | 25 | 2 | 8 |
| D2 | 87 | 12 | 14 |
| Calcitriol | 7 | 2 | 29 |
| Total | 428 | 63 | 15 |
| Fisher's exact test p = 0.399 |  |  |  |

**Table 4.** Bivariate analysis of most recent active vitamin D treatment before most recent vitamin D level at least 14 days before COVID-19 test order and whether that level was <20 ng/ml, among patients with a diagnosis of vitamin D deficiency within 2 years or a level <20 ng/ml.

| Most recent active vitamin D treatment before most recent vitamin D level | Total patients | Vitamin D Deficient (Most recent vitamin D in past year <20 ng/ml) |  |
| --- | --- | --- | --- |
|  |  | N | % |
| No Vitamin D | 139 | 87 | 63 |
| 1-1000 IU D3/Multivitamin | 92 | 50 | 54 |
| 2000 IU D3 | 38 | 13 | 34 |
| 3000+ IU D3 | 25 | 15 | 21 |
| D2 | 107 | 73 | 68 |
| Calcitriol | 5 | 3 | 60 |
| Total | 406 | 241 | 59 |
| Fisher's exact test p = 0.009 |  |  |  |

\*Shown are the last vitamin D levels between 1 year before and 14 days before COVID-19 test orders (N=489 distinct patients). Seven levels are in pg/ml from a test of 1,25(OH)<sub>2</sub>-vitamin D and the remainder are in ng/ml from a test of 25(OH)-vitamin D.

## Discussion

This study provides the first direct evidence of the association of vitamin D deficiency and potentially insufficient treatment with testing positive for COVID-19. The multivariable analysis suggests that persons with deficient most recent vitamin D levels whose treatments were not increased and therefore likely remained vitamin D deficient were at substantially higher risk of testing positive for COVID-19 than were persons with levels that were likely to be sufficient.

This result supports the idea that having an insufficient vitamin D level increases the risk for COVID-19 and suggests that treatment for vitamin D deficiency can reduce that risk. The potential value of treatment for vitamin D deficiency in reducing the risk of COVID-19 is supported by the finding that patients with deficient last vitamin D levels who did have increased treatment were not found to have increased risk for COVID-19 compared to patients whose last vitamin D levels and treatments after those levels suggested they were likely to have had sufficient levels of vitamin D at the time they were tested for COVID-19. Though the difference in COVID-19 risk among these patients with deficient last vitamin D levels who were and were not treated did not reach statistical significance, the small fraction of those receiving treatment who received high dose vitamin D3 suggests that the ability identify an association of treatment in this sample is limited by relatively conservative treatment approaches used. That vitamin D treatment type and dose are related to rates of deficiency and that this association is fully accounted for by the two most effective treatments for vitamin D deficiency studied (vitamin D3 2000IU or ≥3000IU) reinforces the potential value of treatment.

Nevertheless, the complex relationship of treatments to COVID-19 risk and vitamin D levels and deficiency highlights the need for randomized trials to determine whether vitamin D treatment might affect the risk of COVID-19. Testing of vitamin D levels may be an important tool in guiding treatments, and the availability of low-cost home testing for vitamin D may be especially valuable given the benefits of social distancing in COVID-19. However, because vitamin D dosing of up to at least 4000IU or 5000IU is generally considered safe,^23^ taking 4000-5000IU of vitamin D daily may be a reasonable approach for persons without known contraindications to vitamin D supplementation.

Although this appears to be the first data directly connecting vitamin D deficiency and the risk of COVID-19, the idea that adequate vitamin D levels could prevent COVID-19 is supported by multiple trials and meta-analysis that find vitamin D treatment can reduce other viral respiratory infections, among which coronaviruses are common causative organisms. While these benefits would be expected to be greatest among vitamin D deficient persons, the low risks of vitamin D supplementation, the costs and limited access to testing and known seasonality of vitamin D levels all argue for population-level supplementation efforts and perhaps targeted testing for groups at high risk for vitamin D deficiency and/or COVID-19, as noted above. Vitamin D deficiency is influenced by genetic factors, including in African Americans,^24^ so Vitamin D testing and treatment may play an important role in reducing incidence among the African American community where COVID-19 has been particularly prevalent and morbid. This also implies that some risk for COVID-19 might be heritable, which might inform COVID-related decision making for persons with biological relatives who have been affected.

If the incidence of COVID-19 can be reduced by treatment with vitamin D, it is tempting to consider whether vitamin D might reduce COVID-19 transmission. However, caution is required because of the potential importance of asymptomatic persons in COVID-19 spread. Vitamin D is understood to modulate immune function through effects on dendritic cells and T cells^25^, which may promote viral clearance and/or reduce inflammatory responses that produce symptoms. Higher vitamin D levels correlate with lower IL-6 levels, which is notable because IL-6 blocking agents are a major target for controlling cytokine storm in COVID-19.^26,27^ Vitamin D may also affect metabolism of zinc^28^, which has been found to decrease the replication of coronaviruses.^29^ To the extent it decreases viral replication or accelerates viral clearance, vitamin D treatment might reduce spread. On the other hand, if vitamin D reduces inflammation, it might increase asymptomatic carriage and decrease symptomatic presentations, including cough, making it hard to predict its effect on viral spread.

Important limitations must be noted. First, the associations observed may not reflect causal effects of vitamin D deficiency on COVID-19, especially since vitamin D deficiency can reflect a range of chronic health conditions or behavioral factors that plausibly reduce the likelihood of treatment of vitamin D deficiency and increase COVID-19 risk. Nevertheless, the results are robust to including a broad set of demographic and comorbidity indicators. Our analysis also included patients treated with vitamin D2 or calcitriol, and included calcitriol levels in our definition of vitamin D deficiency, which are often used in patients with chronic kidney disease or hypoparathyroidism. In sensitivity analysis, our results generally strengthened when we omitted these patients. We also note our sample is enriched in persons with vitamin D deficiency because of the large number of African Americans, adults with chronic illness and health care workers, all living in a northern city and exposed to COVID-19 during winter.

Vitamin D deficiency may be a smaller risk factor in other populations. The relative simplicity of the analysis performed here would facilitate replication of this analysis in other settings. Since vitamin D deficiency is so prevalent in the US population, and especially in persons with darker skin, who are older and nursing home residents, who are at increased risk for COVID, it seems likely that these findings might be replicated in other settings.

The data presented here argues strongly for a role of vitamin D deficiency in COVID-19 risk and for expanded population-level vitamin D treatment and testing and assessment of the effects of those interventions. Studies of whether interventions to reduce vitamin D deficiency could reduce COVID-19 incidence should be an important priority, including both broad population interventions and intervention among groups at increased risk of vitamin D deficiency and/or COVID-19.

## Data Availability

This data cannot be made publically available due to human subjects issues given the sample size and known location and time frame.

## Funding, Disclosures and Acknowledgements

Meltzer, Best and Zhang wish to acknowledge support from the Learning Health Care System Core of the University of Chicago/Rush University Institute for Translational Medicine (ITM) Clinical and Translational Science Award (ITM 2.0: Advancing Translational Science in Metropolitan Chicago, UL1TR002389, Solway, Contact PI)) and the African American Cardiovascular pharmacogenetic CONsorTium (ACCOuNT, U54-MD010723, Perera, Meltzer Co-PIs). We would like to acknowledge Dr. Stephen Weber for assistance in the UCM operational analyses that informed design of this study, and Tim Filarski and Steven Hooper, who helped with laboratory data acquisition.

## Methodologic Supplement

### Chronic condition indicators

Each chronic condition indicator was set equal to 1 if the patient had at least one of a list of ICD-10-CM diagnosis codes included on an administrative/billing record with a discharge date between January 1, 2018 and April 10, 2020. The lists of codes used for the indicators was defined using the HCUP Elixhauser Comorbidity Software.^20^ For each condition, we used all ICD-10-CM and DRG codes listed within categories in the Elixhauser file comformat_icd10_cm_2020_1.sas, entitled Creation of Format Library for Comorbidity Groups, ICD-10-CM Elixhauser Comorbidity Software, Version 2020.1. The actual ICD-10-CM codes can be found by finding the following category variable names in the file. Hypertension: HTN, HHRWCHF, HHRWHRF, HHRWOHRF, HHRWRF, HRENWORF, HRENWRF, HTNCX, HTNPREG, HTNWCHF, HTNWOCHF, OHTNPREG, HTNCXDRG, HTNDRG. Diabetes: DM, DMCX, DIABDRG. Chronic pulmonary disease: CHRNLUNG, PULMDRG. Pulmonary circulation disorders: PULMCIRC. Depression: DEPRESS. CKD: RENLFAIL, HHRWCHF, HHRWHRF, HHRWOHRF, HHRWRF, HRENWRF, RENALDRG, RENFDRG. Liver disease: LIVER. CM_IS: AIDS, METS, TUMOR, ARTH.

**Supplemental Table 1:**
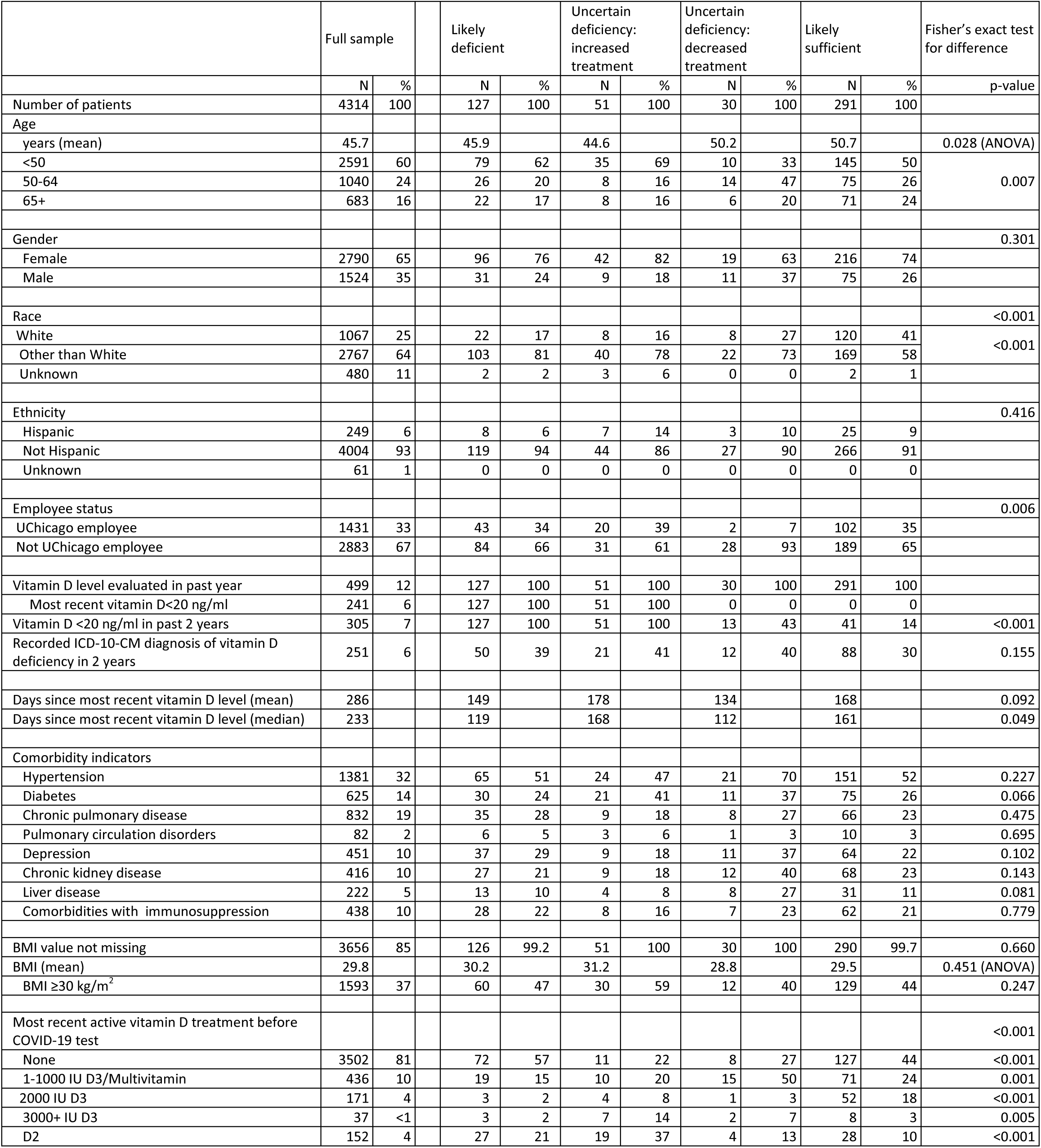
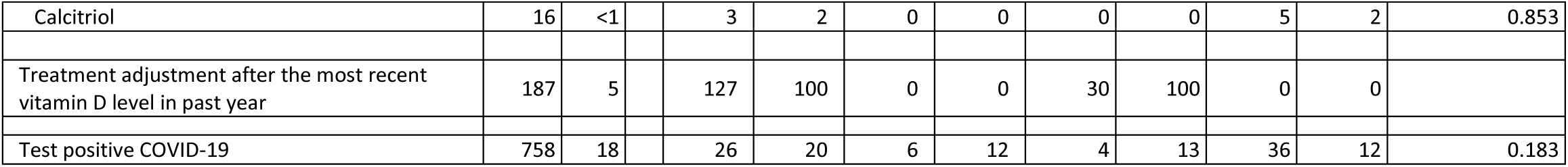
Characteristics of Patient Population

